# K-Edge Imaging Using a Clinical Dual-Source Photon-Counting CT System

**DOI:** 10.1101/2025.08.21.25333798

**Authors:** Martin V. Rybertt, Leening P. Liu, Pooyan Sahbaee, Tristan Nowak, Manoj Mathew, Harold I. Litt, David P. Cormode, Ali H. Dhanaliwala, Peter B. Noël

## Abstract

**Purpose:** To evaluate the feasibility and performance of K-edge imaging of iodine (I) and gadolinium (Gd) on a clinically available photon-counting computed tomography (PCCT) system.

**Methods:** A dual-source clinical PCCT scanner with four energy thresholds (20, 55, 72, 90 keV) was used to scan phantoms containing pure and mixed solutions of I and Gd across multiple concentrations (1–10 mg/mL) and radiation doses (1–8 mGy). Multi-material decomposition was performed using a calibration-based, image-domain algorithm to generate material-specific maps. Quantitative accuracy was assessed using Bland-Altman analysis and contrast-to-noise ratio (CNR), while noise and bias trends were statistically analyzed using non-parametric tests.

**Results:** K-edge imaging was successfully achieved on a clinical PCCT system with accurate decomposition of I and Gd across varying concentrations, solution types (pure/mixed), and dose levels. Quantitative bias was significantly influenced by radiation dose, concentration, and solution type (p < 0.0004). Increased radiation dose and contrast concentration improved quantification accuracy, with maximum bias reductions of 0.9 (I) and 0.3 mg/mL (Gd). CNR correlated linearly with concentration (R² > 0.99) and moderately with dose (R² = 0.85–0.94), achieving peak values of 13 (I) and 16 (Gd) at 8 mGy. Mixed solutions showed reduced performance compared to pure solutions, i.e., CNR of 5 mg/mL Gd solutions increased by 0.6 per mGy in pure solutions while by 0.5 per mGy in mixtures. Noise was dependent on dose but not on concentration or solution type.

**Conclusion:** This study establishes the feasibility of K-edge imaging using a clinical PCCT system and demonstrates accurate, simultaneous decomposition of I and Gd in pure and mixed solutions. These findings support the clinical translation of K-edge imaging and highlight PCCT’s potential for advanced dual-contrast and molecular imaging applications.

## Introduction

Photon-counting computed tomography (PCCT) has emerged as a transformative imaging modality, redefining the capabilities of CT systems.^1–3^ By utilizing photon-counting detectors (PCDs), PCCT measures the energy of individual photons, enabling advanced spectral imaging applications such as K-edge imaging—a technique that exploits the unique X-ray absorption properties of elements with high atomic numbers (Z) to achieve material-specific contrast. Specifically, this technique leverages the K-edge effect to perform material decomposition. This effect refers to the sudden increase in X-ray attenuation when the energy of incoming photons matches or exceeds the binding energy of K-shell electrons in an atom. In contrast, current material decomposition techniques utilize two basis functions to identify materials, commonly the photoelectric effect and Compton scattering. However, employing two basis functions to perform K-edge imaging is insufficient, as these cannot describe the discontinuity in the attenuation profile of K-edge materials. As a result, dual-energy techniques alone are insufficient for capturing this effect unless they incorporate assumptions or leverage prior knowledge. Presently, clinically available PCCT systems can acquire spectral information with up to four energy thresholds, facilitating K-edge imaging.

K-edge imaging has been extensively studied over the past decades for applications ranging from material decomposition techniques to quantitative imaging.^4–7^ Currently, K-edge imaging research has shifted its focus to its translation into clinical practice in parallel to assessing the viability of potential K-edge materials. Because the human body naturally lacks elements with significant K-edge signals in the 40-90 keV energy range. K-edge utility comes from contrast agents, allowing for contrast enhancement of the vasculature and tissue. Among high-Z elements with K-edges within this range, gadolinium (Gd) is especially significant due to its dual role in medical imaging. It is widely utilized in magnetic resonance imaging (MRI) and has been employed in CT as an alternative contrast agent (CA) for patients who cannot tolerate iodinated CAs,^8^ as demonstrated in conventional CT angiography.^9,10^ While Gd has a higher Z (64) and K-edge (50.2 keV) than iodine (I) (Z: 53, K-edge: 33.2 keV), the resulting contrast of Gd on conventional CT is limited by its small maximum allowed dose (0.2 mmol/kg). Conversely, iodine is a poor K-edge material for diagnostic CT because the photon flux on either side of its K-edge is unbalanced, producing a negligible K-edge signal. Although PCCT and dual-energy CT (DECT) can reconstruct virtual monoenergetic images on both sides of a K-edge, the underlying models do not account for K-edge effects. Consequently, evidence from phantom studies,^11–15^ animal models,^16,17^ and clinical research^18,19^ suggests that the overall diagnostic gain remains limited.

Compared to DECT, PCCT systems incorporate energy resolution via multi-bin imaging and sophisticated material decomposition algorithms, making the generation and application of K-edge-specific material maps feasible.^20,21^ Extensive research on K-edge imaging applications has been conducted using prototype PCCT systems. A particularly promising application is dual-contrast imaging, which enables the simultaneous visualization of characteristic arterial and portal venous enhancement within a single scan, particularly for liver imaging.^22^ This approach reduces the need for sequential scans, lowers radiation exposure, and enables a broad range of clinical applications, particularly in complex scenarios such as oncology and vascular imaging. In conjunction with the development of prototype PCCT systems, dual-contrast protocols — primarily with I and Gd—were evaluated through simulation and phantom studies^23–28^ as well as cardiovascular and abdominal applications in animal models.^22,29–34^ Furthermore, in vivo studies not only demonstrated the capability of differentiating I and Gd, but also the significant potential to separate bone/calcium from CAs via K-edge imaging.^35,36^ Ongoing research focuses on developing novel contrast agents, including high-Z nanoparticles with K-edges in the diagnostic energy range.^37,38^

Despite the demonstrated advantages of PCCT, practical implementation of K-edge imaging has remained out of reach. This study addresses an important gap in the translation of K-edge imaging into clinical practice by evaluating Gd K-edge imaging with a clinically available PCCT system. We illustrate the current capabilities of clinical PCCT systems to perform K-edge imaging of Gd via a phantom study to examine the relationship between quantitative accuracy and image quality to contrast concentration, radiation dose, and type of solution.

## Methods

### Material decomposition

To evaluate the feasibility of K-edge imaging, we utilized a clinically approved PCCT system (NAEOTOM Alpha, Siemens Healthineers, Forchheim, Germany) capable of multi-material decomposition (MMD). It enables the separation of photons into four distinct energy bins, allowing for the generation of up to four material maps. Neumann et al.^20^ describes the MMD approach used in this study to perform K-edge imaging. This image-based algorithm is a prior knowledge-free approach that calibrates each material of interest based on measurements derived from the four energy bins. During calibration, a set of expected values is produced to serve as ground truth for later estimation steps. In the MMD process, input images are initially corrected for water, followed by material decomposition. Then, least squares estimation is performed using the calibration data as a reference, resulting in material-specific maps for each target material.

### Calibration

The MMD calibration process involved repeated scanning for each target material while varying object diameter, tube current, and the distance from the object to the isocenter. We prepared water, I (13.5 mg/mL), and Gd (10.0 mg/mL) solutions and placed them inside a 10 cm diameter solid water-equivalent phantom provided by the vendor (Siemens Healthineers, Forchheim, Germany) (Figure 1 A). The solutions were then scanned at varying tube currents and isocenter distances, with 10 cm extension rings added stepwise to increase the diameter up to 40 cm (Figure 1 B).

**Figure 1.**
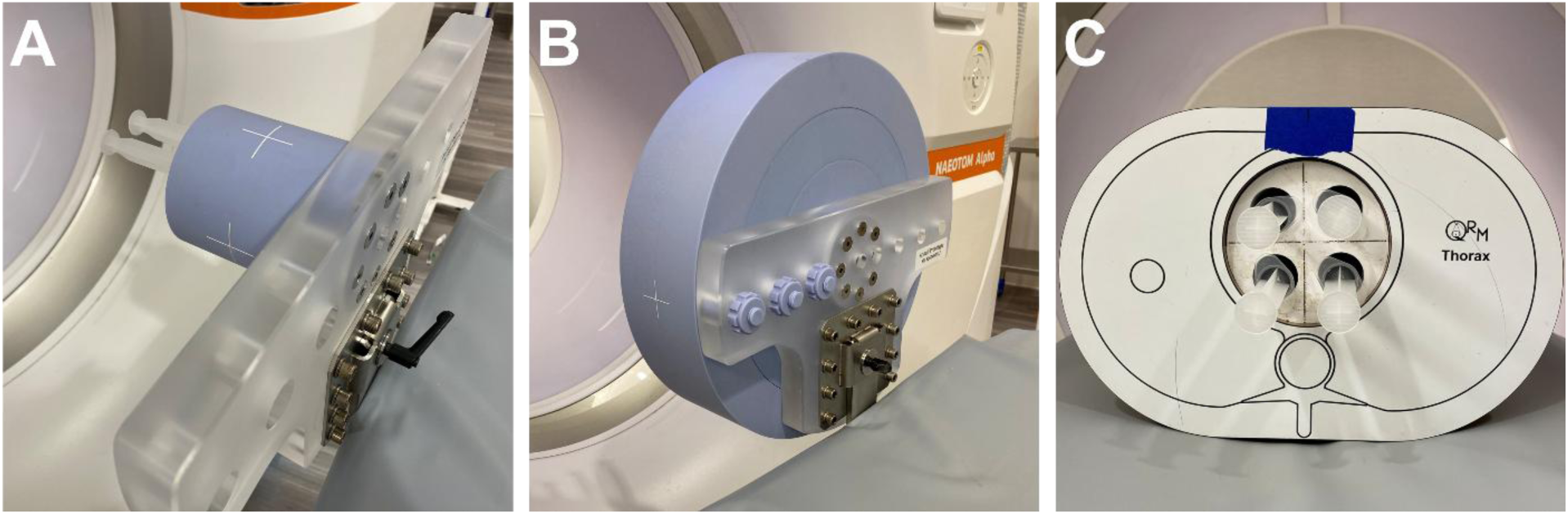
Calibration and experimental setup. The calibration phantom (**A**), containing a syringe filled with the calibration material (water, I, or Gd) and water-mimicking inserts, was scanned with a dual-source photon-counting CT. Additional 10 cm extension rings (**B**) were added after completion of each set of calibration scans. To evaluate the feasibility and accuracy of K-edge imaging, syringes filled with contrast agent solutions were placed within a thoracic phantom and scanned (**C**).

### Experimental setup

Solutions comprised of either I (Isovue-300, Bracco Diagnostics, Milan, Italy) or Gd (Dotarem, Guerbet LLC, Princeton, United States) were prepared at concentrations of 1, 2.5, 5, and 10 mg/mL (pure solutions). To assess whether the separation of I and Gd was feasible within the same volume, mixed solutions were prepared with ratios of 1:2.5, 2.5:1, 2.5:2.5, 2.5:5, 5:2.5, and 5:5 mg/mL of I and Gd, respectively. Solutions were placed in a 10 cm diameter solid water-equivalent holder that was fitted into a thoracic phantom (QRM Thorax, QRM, Möhrendorf, Germany) (Figure 1 C) for scanning.

### Image acquisition

A dual-source clinical PCCT system was used to scan the phantom setup in an acquisition mode that utilizes energy thresholds at 20, 55, 72, and 90 keV, a tube voltage of 140 kVp, and a volumetric CT dose index (CTDI_vol_) of 1, 2, 4, and 8 mGy (Table 1). Each scan was repeated three times for reproducibility, with raw data collected for subsequent processing on a vendor-specific offline workstation.

**Table 1.**
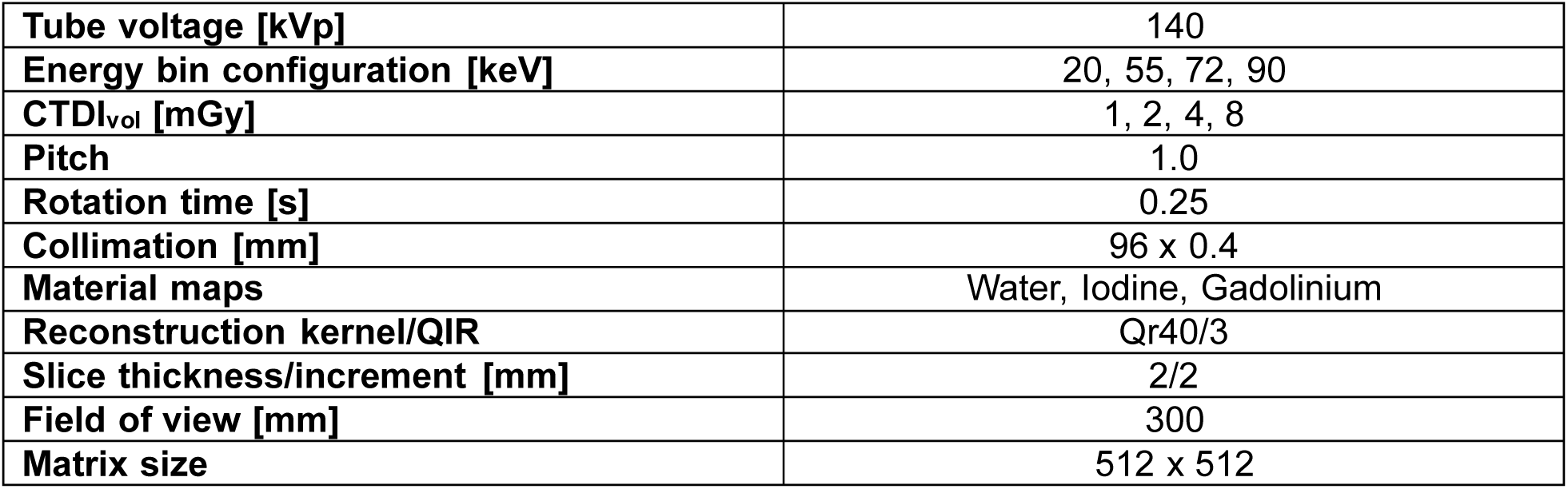
Acquisition and reconstruction parameters.

|  |  |
| --- | --- |
| **Tube voltage [kVp]** | 140 |
| **Energy bin configuration [keV]** | 20, 55, 72, 90 |
| **CTDI<sub>vol</sub> [mGy]** | 1, 2, 4, 8 |
| **Pitch** | 1.0 |
| **Rotation time [s]** | 0.25 |
| **Collimation [mm]** | 96 x 0.4 |
| **Material maps** | Water, Iodine, Gadolinium |
| **Reconstruction kernel/QIR** | Qr40/3 |
| **Slice thickness/increment [mm]** | 2/2 |
| **Field of view [mm]** | 300 |
| **Matrix size** | 512 x 512 |

### Image reconstruction

Scans were reconstructed into specialized spectral post-processing (SPP) files using a proprietary workstation (ReconCT, Siemens Healthineers, Forchheim, Germany) at a field of view (FOV) of 300 mm, pixel spacing of 0.59x0.59 mm, using a Qr40 filter, and quantum iterative reconstruction (QIR) level of 3. Consequently, material decomposition was performed for each material of interest (water, iodine, and gadolinium) using vendor-specific software (eXamine, Siemens Healthineers, Forchheim, Germany).

### Image analysis

Regions of interest (ROIs) were placed at the center of each sample solution, covering 60% of the diameter of the samples to measure mean concentration and standard deviation across 15 central slices, as well as at the center of the water-equivalent holder to measure background noise. This information was collected and used to perform a Bland-Altman analysis to assess quantification accuracy, as well as to calculate the contrast-to-noise ratio (CNR) to evaluate image quality and impact of noise. Due to the lack of true reference values, measurements obtained at 8 mGy were treated as a baseline to compute relative differences, serving as an indirect measure of bias.

### Statistical analysis

Statistical analyses were carried out in Python using non-parametric methods appropriate for grouped data. Measured concentrations, image noise, and difference relative to reference bias were categorized by material. Variables with two groups (material and solution type) were tested using the Mann-Whitney U test, while those with multiple groups (concentration and radiation dose) were analyzed with the Kruskal-Wallis test, followed by Dunn’s post hoc test to identify specific group differences. To control the risk of false positives across multiple comparisons (n = 117), the significance threshold was adjusted using the Bonferroni correction, resulting in a p-value cutoff of 0.0004.

## Results

### Material-specific images

The feasibility of K-edge imaging on a clinically available PCCT system is demonstrated in **Figure 2**. Material-specific imaging of iodine and gadolinium achieved accurate decomposition and quantification, with sufficient contrast observed even in the absence of additional denoising. Contrast levels corresponded well with expected concentrations; however, in mixtures with unequal concentrations, a slight bias was noted toward the material present in higher concentration.

**Figure 2.**
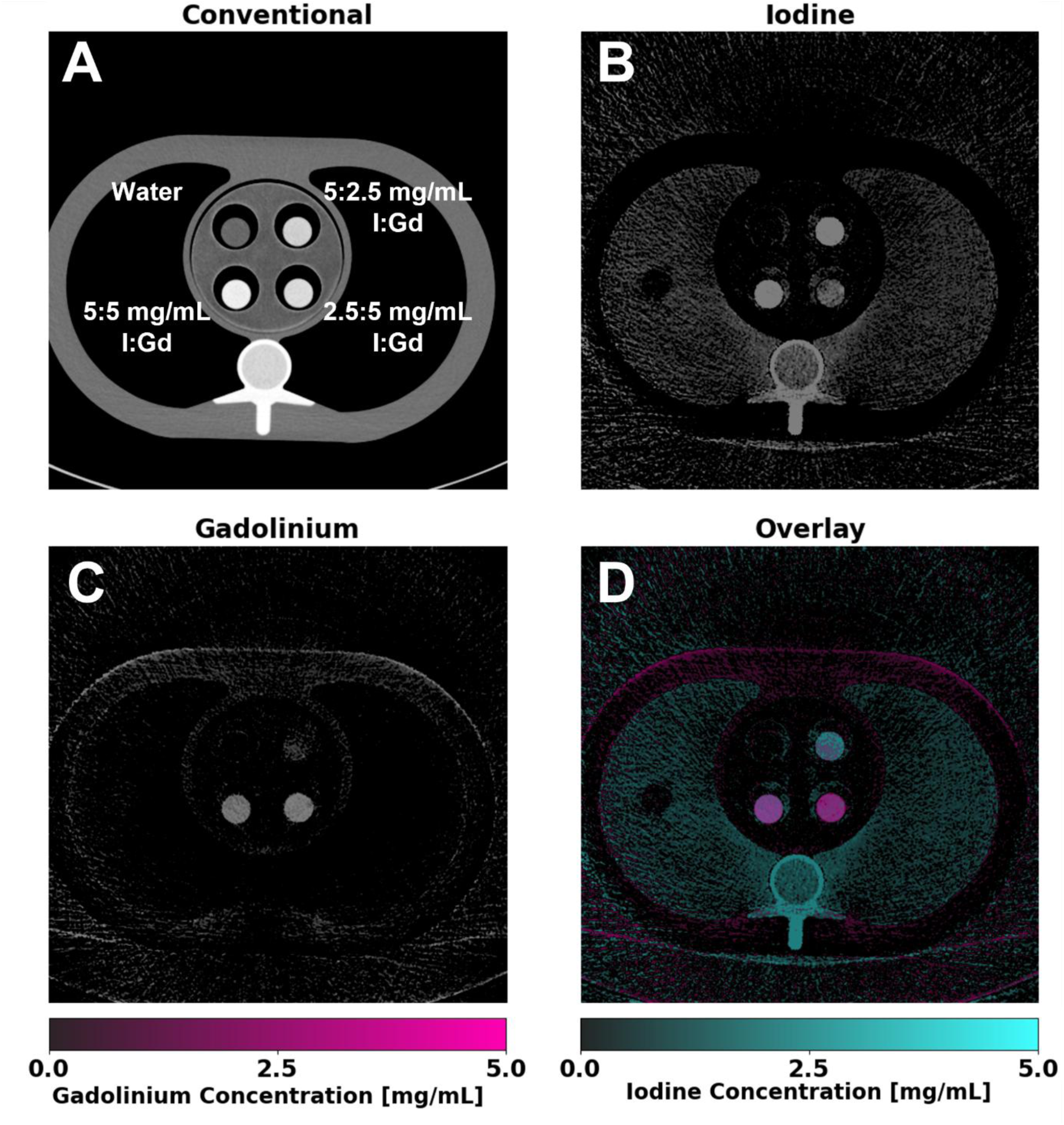
Material-specific images of gadolinium and iodine using K-edge imaging. Inserts containing varying concentrations of I and Gd were placed within a thoracic phantom and scanned. Conventional images (**A**) display a standard grayscale CT attenuation image (WL/WW: 200/600 HU), illustrating the inability to separate the two contrast agents. Iodine maps (**B**) (WL/WW: 2.5/5 mg/mL) show successful localization of iodine within inserts, with gadolinium maps (**C**) (WL/WW: 2.5/5 mg/mL) also achieving successful separation of gadolinium and iodine within the same volume. An overlay image (**D**) highlights the spatial distribution of both contrast agents, enabling enhanced visualization of co-localized and distinct regions of each contrast agent.

### Quantitative accuracy

K-edge imaging of I and Gd demonstrated high accuracy and consistent material decomposition across varying contrast concentrations, solution types, and radiation dose levels. Bland-Altman analysis revealed only minor inaccuracies, with narrow confidence intervals observed across all parameters (Figure 3). The difference from the reference was significantly influenced by dose, concentration, and solution type (p < 0.0004). Specifically, increasing the dose from 1 to 4 mGy led to a significant improvement in accuracy. For pure solutions, bias was reduced by 0.2 ± 0.4 mg/mL I and 0.2 ± 0.3 mg/mL Gd. Mixed solutions showed a similar trend, with bias reductions of 0.1 ± 0.2 mg/mL I and 0.2 ± 0.2 mg/mL Gd. Likewise, contrast concentration had a significant effect (p < 0.0004) across both pure and mixed solutions. As concentration increased from 1 to 5 mg/mL, pure solutions showed a reduction in bias of 0.9 mg/mL I and 0.3 mg/mL Gd, while mixed solutions showed reductions of 0.4 mg/mL I and 0.3 mg/mL Gd. A detailed summary of these results is provided in Table 2.

**Figure 3.**
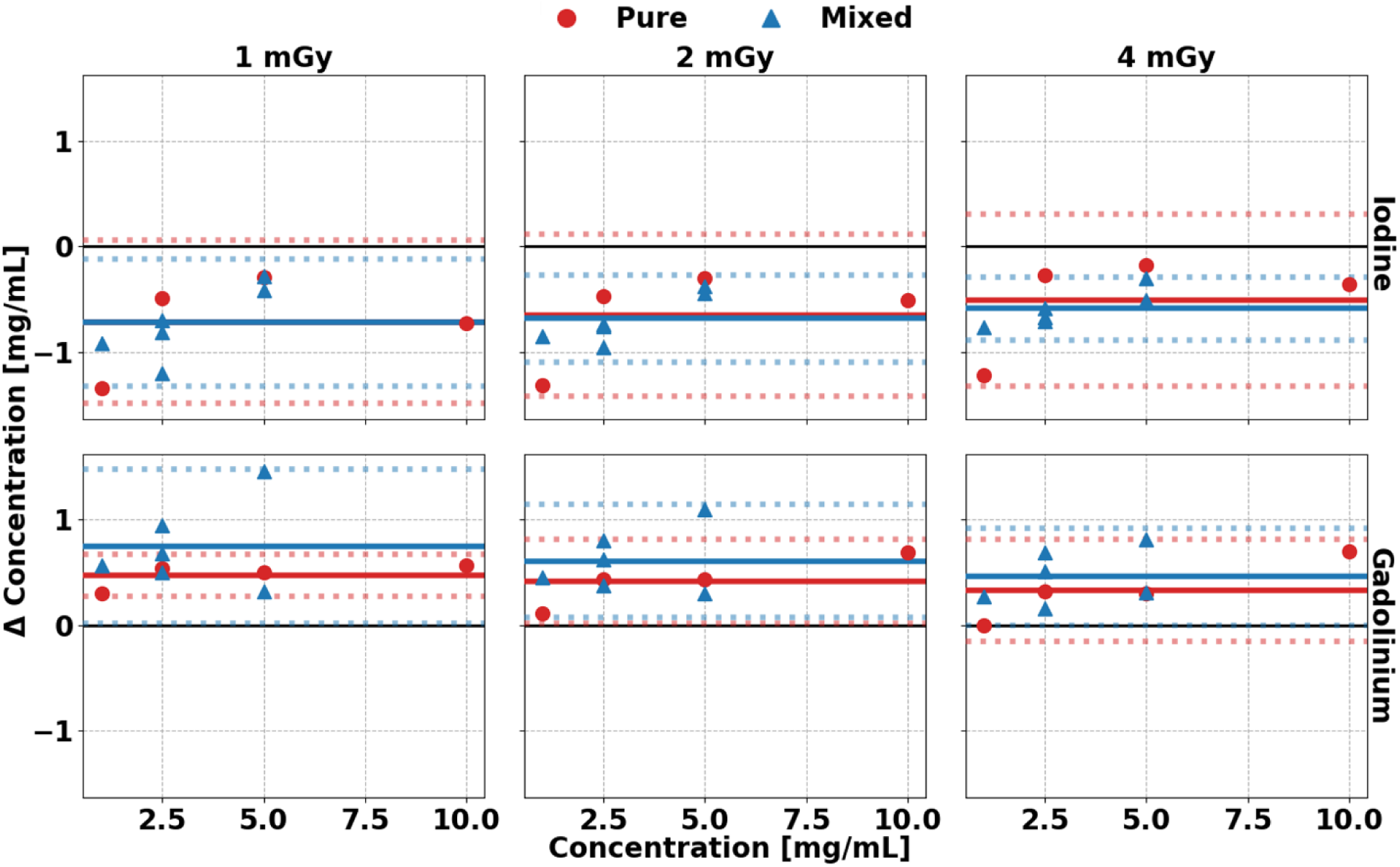
Bland-Altman plot of iodine and gadolinium material maps at each radiation dose. Material decomposition of I (**A**) and Gd (**B**) demonstrated stable quantification across different concentrations, types of solutions, and radiation doses for all parameters. Solid lines represent the mean difference between measured and expected concentration, whereas dashed lines represent the 95^th^ percentile confidence interval.

**Table 2.** Bias on I and Gd-specific images relative to measured concentration on 8 mGy images.

| CTDI<sub>vol</sub><br>[mGy] | I |  | Gd |  |
| --- | --- | --- | --- | --- |
|  | Bias [mg/mL] |  | Bias [mg/mL] |  |
|  | Pure<br>solution | Mixed<br>solution | Pure<br>solution | Mixed<br>solution |
| 1 | $-0.7 \pm 0.4$ | $-0.7 \pm 0.3$ | $0.5 \pm 0.3$ | $0.8 \pm 0.4$ |
| 2 | $-0.7 \pm 0.4$ | $-0.7 \pm 0.3$ | $0.4 \pm 0.3$ | $0.6 \pm 0.3$ |
| 4 | $-0.5 \pm 0.4$ | $-0.6 \pm 0.2$ | $0.3 \pm 0.3$ | $0.4 \pm 0.3$ |

### Contrast and noise

Material decomposition of I and Gd exhibited considerable contrast and demonstrated significant detectability. Notably, a maximum CNR of 13 and 16 was observed for I and Gd, respectively, in pure solutions at their highest concentration and dose (Figure 4). A linear regression revealed a strong correlation between CNR and concentration, where CNR at a radiation dose of 8 mGy increased by 1.3 (R^2^ = 0.9930) and 1.6 (R^2^ = 0.9995) per mg/mL for I and Gd, respectively. Similarly, CNR in mixed solutions was comparable (Figure 5), demonstrating an increase of 1.3 (R^2^ = 0.9943) and 1.6 (R^2^ = 0.9742) per mg/mL of I and Gd, respectively, while also reaching a peak CNR of 7 and 7.2. In contrast, CNR exhibited a linear correlation with radiation dose with lower correlation coefficients, demonstrating a smaller effect of dose on detectability. CNR increased by 0.4 (R^2^ = 0.8460) and 0.6 (R^2^ = 0.9377) for each 1 mGy for pure solutions of I and Gd at 5 mg/mL, respectively, while mixed solutions produced similar slopes of 0.4 (R^2^ = 0.8854) and 0.5 (R^2^ = 0.8454) at a concentration of 5 mg/mL. Noise in material-specific images of I and Gd only revealed a significant dependence (p < 0.0004) on radiation dose, whereas concentration and solution type did not present a significant effect on noise. Noise in I-specific images did not vary between pure and mixed solutions, with an average noise being equal across radiation doses at 1.5 ± 0.02, 1.1 ± 0.01, 0.8 ± 0.01, and 0.6 ± 0.01 mg/mL for CTDI_vol_ of 1, 2, 4, and 8 mGy, respectively. In comparison, Gd-specific maps show the same trend, where noise was 1.5 ± 0.02, 1.0 ± 0.01, 0.8 ± 0.01, and 0.6 ± 0.01 mg/mL across the same range of radiation doses.

**Figure 4.**
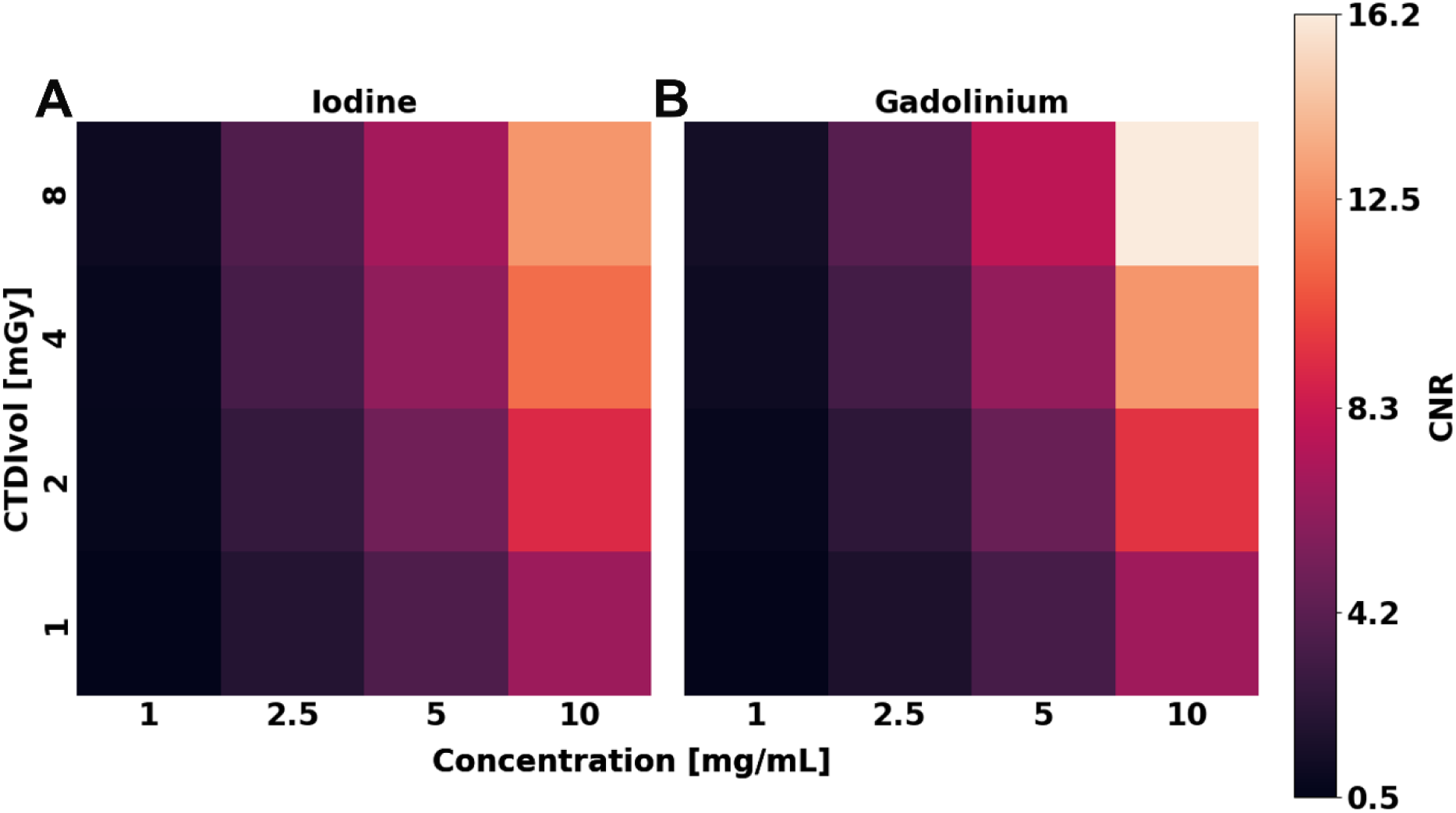
Contrast-to-noise ratio of iodine and gadolinium in pure solutions at different concentrations and radiation doses. Higher concentration of material and radiation dose improved contrast in I (**A**) and Gd (**B**) material maps. CNR presented an approximately ten-fold increase between the lowest and highest concentration, and a two-fold increase between the lowest and highest radiation dose.

**Figure 5.**
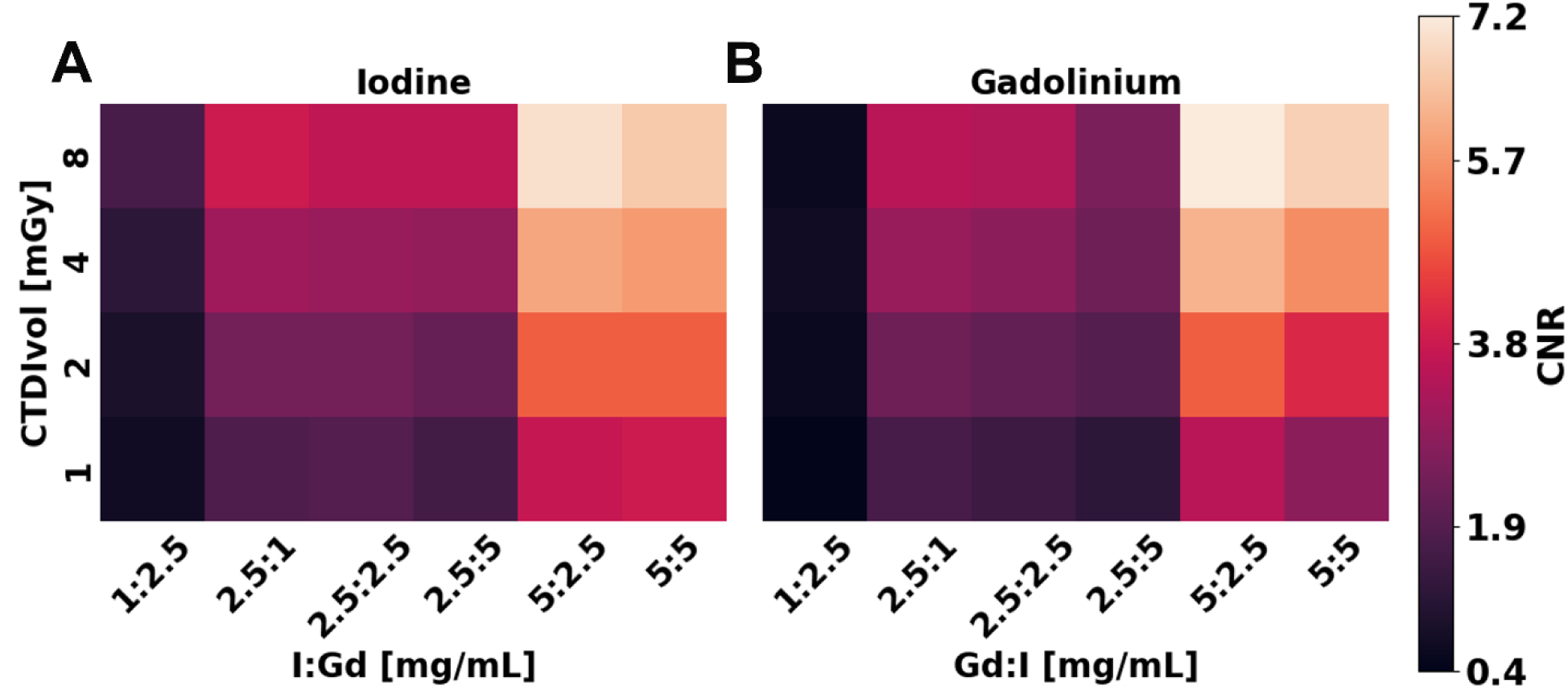
Effect of radiation dose and contrast agent concentration on contrast of material decomposition maps. I (**A**) and Gd (**B**) CNR were primarily influenced by contrast agent concentration, while radiation dose rendered a smaller effect on CNR. CNR was higher for mixed solutions where the concentration of one CA corresponding to the material map was greater than the other CA, i.e., CNR at 8 mGy of 5:2.5 mg/mL I:Gd is 7.9 while the 5:5 solution is lower at 7.1.

## Discussion

This study demonstrated the feasibility of Gd K-edge imaging using a clinical dual-source PCCT system, marking a key step toward integrating advanced spectral imaging into routine clinical workflows. Leveraging PCCT’s superior energy discrimination capabilities, we achieved simultaneous, material-specific imaging of I and Gd within a single acquisition. Our findings confirmed accurate separation and quantification of both CAs, underscoring PCCT’s potential for dual-contrast imaging applications.

PCCT systems are capable of multi-energy acquisitions, thus overcoming the limitations of DECT systems for K-edge imaging—the restriction of acquiring only two sets of spectral information. In this study, we demonstrated the capability of a clinical PCCT system to perform K-edge imaging of relevant CA materials such as Gd. Quantitative performance was significantly influenced by radiation dose, contrast concentration, and solution type, with higher doses and concentrations reducing the bias relative to the reference. Compared to pre-clinical PCCT studies of K-edge imaging,^34^ which reported biases of 0.75 mg/mL for iodine and 0.45 mg/mL for gadolinium, our results demonstrated comparable or improved accuracy. Estimated biases ranged from 0.5 to 0.7 mg/mL I and 0.3 to 0.7 mg/mL Gd, achieved using lower radiation doses. Prior dual-contrast imaging studies using DECT relied on assumptions such as volume conservation to perform K-edge imaging, allowing systems that are unable to acquire multi-energy acquisitions to perform material decomposition. In comparison, these show improved accuracy to our study, exhibiting errors ranging from 0.1 to 0.7 mg/mL in decomposition of Gd.^11,12^ These systems often suffer from greater bias and diminished image quality when their underlying assumptions break down, as it commonly occurs in an everyday clinical context. Our method offers an alternative: assumption-free K-edge imaging using a clinical PCCT platform. Moreover, this approach opens the door to the use of novel contrast agents— including gold, ytterbium, and tungsten^39–41^ —which offer improved targeting, prolonged circulation times, and lower biotoxicity but require quantitative K-edge detection for clinical viability. Overall, our results emphasize the transformative potential of PCCT in enabling precise, quantitative decomposition of multiple contrast agents, supporting the next generation of functional and molecular CT imaging.

Despite the complex task of separating mixtures of CAs within the same voxel, we demonstrated consistent image quality of I and Gd-specific images across solution types. We observed that the decomposition of pure solutions outperforms that of mixed solutions. Specifically, the CNR of mixed solutions at the same radiation dose and concentration was smaller than that of pure solutions. Nevertheless, our findings display an expected increase in CNR due to increased dose and concentration^42^ across both solution types. A previous study, utilizing a pre-clinical system, similarly reported a lower performance in mixed solutions compared to pure solutions.^25^ However, they also described a higher performance in comparison to our study, showing an increase in CNR of 3.2 and 3.0 per mg/mL of I and Gd, respectively, whereas our estimated CNR increased by 1.3 and 1.6, respectively. The higher performance may in part be due to the use of a smaller phantom, which improves dose efficiency, and possibly also to the use of a projection-based approach, which has in some cases been reported to reduce bias and noise compared to image-based methods.^43^ Additionally, our measured CNR values are in accordance with previous MMD of I and Gd in ex vivo studies and animal models.^29,31,35^ As ex vivo and animal studies present less favorable conditions, this underscores the need for developing additional denoising techniques to improve image quality in K-edge imaging.

This study presents some limitations. Firstly, the approved clinical dosage for Gd-based contrast agents (0.2 mmol/kg) is small and can result in lower contrast enhancement in target areas compared to iodinated CAs in CT. While we evaluated the performance of Gd K-edge imaging with concentrations within the expected range of clinical concentrations from 0.6 to 4.8 mg/mL,^11,12^ this can still result in insufficient contrast that is not fully addressed by the increased detectability of material-specific images. To fully unlock the potential of K-edge imaging, alternative K-edge materials with higher Z could provide improved contrast for K-edge imaging applications. Secondly, MMD introduces higher noise levels compared to two-basis material decomposition. This is primarily due to the division of the x-ray spectrum into additional energy bins, which reduces the number of photons per bin and, consequently, increases input noise during decomposition. As a result, multi-bin decomposition techniques require dedicated denoising strategies to maintain image quality. Some successful strategies for K-edge imaging applications include deep learning approaches embedded in the decomposition process to mitigate the additional input noise.^44,45^ Thirdly, current clinical PCCT systems offer three threshold configurations: (20, 40, 56, 75 keV), (20, 52, 75, 82 keV), and (20, 55, 72, 90 keV). Configuration selection can result in improved performance and has been shown^46^ to be related to the K-edge materials in use. In this study, we selected a configuration that ensured adequate separation from the K-edge of Gd (50.2 keV). However, further optimization is required to determine the ideal bin position for accurate and robust K-edge imaging of Gd.

This study exhibited the clinical feasibility of Gd K-edge imaging using a clinical dual-source PCCT system, enabling accurate decomposition and quantification of iodine and gadolinium within a single acquisition. These results support the integration of PCCT into advanced spectral imaging workflows and highlight its potential for expanding clinical applications through multi-material, quantitative imaging.

## Data Availability

All data produced in the present study are available upon reasonable request to the authors.

## Acknowledgement

We acknowledge support through the National Institutes of Health (R01EB030494, & R01EB035908) and Siemens Healthineers.

## Abbreviation Definition

PCCT: Photon-counting computed tomography
PCD: Photon-counting detector
CT: Computed tomography
MRI: Magnetic resonance imaging
CA: Contrast agent
I: Iodine
Gd: Gadolinium
keV: Kiloelectron volt
Z: Atomic number
MMD: Multi-material decomposition
CTDI_vol_: Volumetric computed tomography dose index
SPP: Spectral post-processing
FOV: Field of view
QIR: Quantum iterative reconstruction
ROI: Region of interest
CNR: Contrast-to-noise ratio
R²: Coefficient of determination (from linear regression)

